# The impact of the undetected COVID-19 cases on its transmission dynamics

**DOI:** 10.1101/2020.05.30.20117838

**Authors:** Sujata Saha, Sumanta Saha

## Abstract

**Objective:** The COVID-19 pandemic is currently ongoing. Presently, due to the unavailability of a definitive vaccine to decrease its acquiring, it’s essential to understand its transmissibility in the community by undetected cases to control its transmission. This study aims to study this context using mathematical modelling.

**Methods:** A COVID-19 transmission model was framed that estimated the basic reproduction number (*R*_0_, a measurement of disease risk) using the next-generation method. It explored the contribution of exposed and infected (detected and undetected) individuals, and environmental pathogen to the overall risk of infection spreading, utilizing the publicly reported data of this infection in Maharashtra between March 22, 2020, and May 4, 2020. A sensitivity analysis was performed to study the effect of a rising number of undetected cases to *R*_0_.

**Results:** The estimated basic reproduction number is ***R*_0_ = 4.63**, which increases rapidly with the rise in the undetected COVID-19 cases. Although the exposed individuals made the largest contribution to infection transmission (***R*_1_ = 2.42**), the contaminated environment also played a significant role.

**Conclusions:** It is crucial to identify the individuals exposed and infected to COVID-19 disease and isolate them to control its transmission. The awareness of the role of fomites in infection transmission is also important in this regard.

## 1. Introduction

During a viral pneumonia outbreak in Wuhan, China, the first case of coronavirus disease 2019 (COVID-19) was reported in December 2019 [1,2]. The causative agent Severe Acute Respiratory Syndrome Coronavirus-2 (SARS-CoV-2), an enveloped single-stranded RNA virus, has a zoonotic origin [2,3]. Similar to Severe Acute Respiratory Syndrome Coronavirus (SARS-CoV) in 2002 and Middle East respiratory syndrome coronavirus (MERS-CoV) in 2012, the symptoms of this infection include dry cough, fatigue, breathing difficulty, and bilateral lung infiltration [3]. In April 2020, Centre for Disease Control (CDC) suggested six additional symptoms of COVID-19: fever, chills, repeated shaking with chills, muscle pain, headache, sore throat, and new loss of taste or smell [4–6].

Since its onset, the disease has been spreading at a very fast pace to different parts of the globe. As of 24 January 2020, 830 confirmed cases had been diagnosed in China, Thailand, Japan, South Korea, Singapore, Vietnam, Taiwan, Nepal, and the United States [7]. Subsequently, the World Health Organization (WHO) formally declared this epidemic as a Global Public Health Emergency of International Concern on January 30, 2020 [3]. The global confirmed case of COVID-19 rose to 266073, with 11184 deaths, as on 22 March 2020 [8].

India reported its first case of COVID-19 on 30th January 2020 [9]. It was found in a Kerala based student who was studying at Wuhan University, China [9]. Soon the disease spread to its other parts and by 22 March 2020, WHO confirmed 360 positive COVID-19 cases across India.[8] Subsequently, on 24th March 2020, a nationwide lockdown was imposed in in India to reduce the pace of COVID-19 spread [10]. Maharashtra is the most affected Indian state which reported 14541 confirm cases including 583 deaths and 2465 recovery as on 4 May 2020 [11,12].

As there is no effective therapeutics or licensed vaccine to combat novel COVID-19, it adds challenges to its control [3,13]. Moreover, as the infected individuals remain asymptomatic during its incubation period (2–14 days), they might keep spreading the disease unknowingly to those in their surroundings [3]. While it is important to test for COVID-19 cases, India is unable to test most of its individuals for new coronavirus cases due to lack of enough test kits, and testing is done in those with symptoms [14]. The nation is combating the disease by tracing people who might have had contact with COVID-19 detected individuals based on their verbal autopsy, and by quarantining them [14]. But, due to the plausible risk of recall bias, some contacts will always remain undetected during such contact tracing exercises. These undetected cases pose a risk of COVID-19 transmission to the community where they live in; therefore, it is crucial to study their role in disease transmission.

Several, research papers published in 2020 have tried to understand the transmission dynamics of this disease by mathematical modelling. Most of these mathematical models studied the disease risk via direct human-to-human transmission route. Choi & Ki [1] framed a susceptible-exposed-infected-hospitalized-recovered (SEIHR) model and calculate the basic reproduction number as 4.028 (95% CI: 4.010,4.046) using the reported number of confirmed cases from January 20, 2020, and March 4, 2020, in Korea. Yang & Wang [3] developed a mathematical model considering the pathogen concentration in the environmental reservoir and its role in disease transmission. In this model, they approximated the overall disease risk *R*_0_ = 4: 25 with *R*_3_ = 1:497 (the infection risk caused by the environmental reservoir of the pathogen), showing a significant contribution from the environmental reservoir toward the overall infection risk. Mandal et al. [13] framed an SEIR model with an objective to prevent, or delay, the local outbreaks in India through restrictions on travel from COVID-19 affected countries. Read et al. [15] reported the basic reproduction number as 3.11 by fitting data of a deterministic SEIR metapopulation transmission model. Li et al. [16] analyzed data on the first 425 confirmed cases in Wuhan and reported the mean incubation period as 5.2 days and the basic reproduction number as 2.2 (95% CI, 1.4 to 3.9). Despite these modelling efforts, the role of undetected cases on the risk of infection spreading in the community remains underexplored.

Therefore, this paper aims to study this under-explored area of COVID-19 infection by mathematical modelling, assuming the disease acquiring potential of susceptible individuals with contact to this virus-infected asymptomatic and symptomatic individuals, and virus-contaminated fomites.

## 2. Notations

We used the following notations to develop the model –

*S* Susceptible individuals
*E* Exposed persons
*I*_1_ Infected people, who are detected through proper testing
*I*_2_ Infected individuals who are undetected
*R* Recovered population
*β_e_* Transmission rates between the exposed and susceptible individuals
*β*_*i*1_ Transmission rates between the detected infected and susceptible individuals
*β*_*i*2_ Transmission rates between the undetected infected and susceptible individuals
*β*_*v*_ Environment to human transmission rates
Π Influx rate in the population
*μ* Natural death rate in the population
*γ* The disease recovery rate
*w* Disease induced death rate
*α*^−1^ Incubation period
*β* Percentage of the infected individuals who are undetected
*ξ*_1_ Contribution of the exposed individuals to the environmental reservoir
*ξ*_2_ Contribution of the detected infected individuals to the environmental reservoir
*ξ*_3_ Contribution of the undetected infected individuals to the environmental reservoir
*σ* Removal rate of the coronavirus from the environment

## 3. Mathematical methods

In this model, we divided the total human population into four compartments – susceptible (S), exposed (E), infected detected (*I*_1_), infected undetected (*I*_1_), and recovered (R). Individuals in the exposed class are asymptomatic infected, but they are capable to spread the disease. People in the both the infected classes (detected and undetected) are fully symptomatic, but *I*_1_ consists those who are detected by proper testing system and are isolated, whereas, *I*_2_ represents the undetected cases who are living in the society as usual. An additional compartment has been introduced for the environmental reservoir of this novel pathogen. The following differentials equations describe the transmission dynamics of the disease COVID-19 –

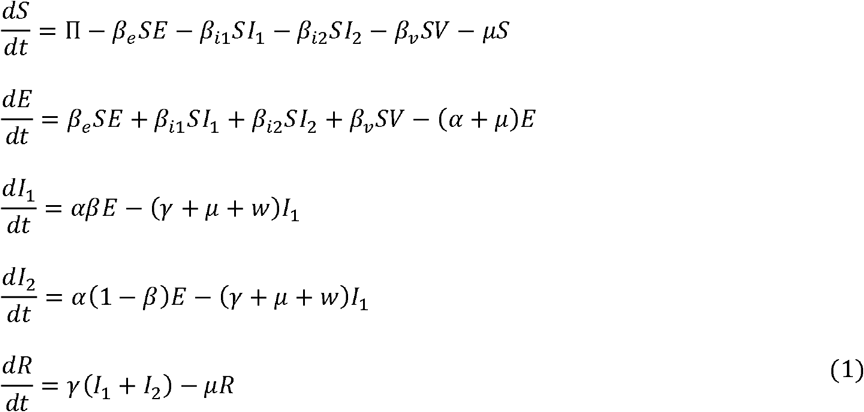

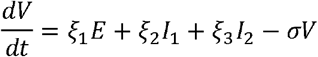

Clearly, the system has a disease-free equilibrium at 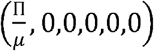.

In this system the disease components are *E, I*_1_, *I*_2_, and *V*. The infection matrix F and the transition matrix V are given by –

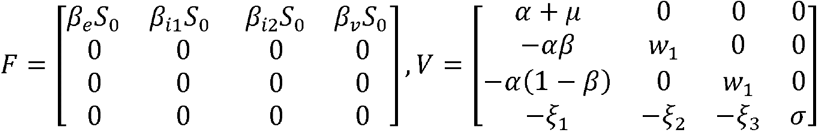

where, 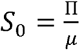, *and w*_1_ = (*γ* + *μ* + *w*)

The basic reproduction number *R*_0_ of the model (1), is defined by the spectral radius of the next-generation matrix *FV*^−1^, therefore,

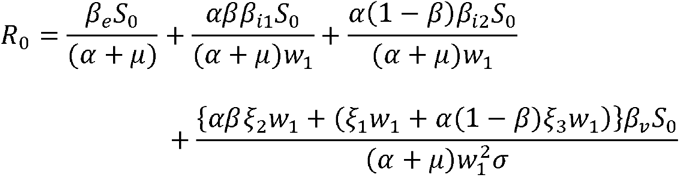

Or

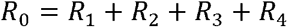

where, *R*_1_, *R*_2_, *R*_3_ and *R*_4_ gives the quantification of the disease risk through susceptible-to-exposed, susceptible-to-infected detected, susceptible-to-infected undetected, and environment-to-human transmission routes, respectively.

## 4. Numerical results

Here, this model is applied to estimate the basic reproduction number using the COVID-19 outbreak data published by WHO and the Ministry of Health and Family Welfare, India for the epidemic period between March 22 and May 4, 2020 for the Indian state of Maharashtra [8,11,12]. The data provides the daily confirm cases, recoveries, and the total death caused by COVID-19.

Since the city was in complete lockdown during this period, its population influx rate was expected to be equal to its birth rate, which was estimated at 5603/day [17]. The total population of Maharashtra (N) is 112374333 [18]. As of 22-March 2020, the total confirmed coronavirus cases were 69 including 2 death and 0 recoveries, but these counts rose to 14541, 583, and 2465 respectively on 4th May 2020 [11,12]. Therefore, *S*(0) = 112374264. Again, as both the exposed and undetected infected persons tend to live in the society, as usual, they transmit the disease at the same rate, the values of these two parameters *β_e_* and *β*_*i*2_ were taken as equal to 0.25 × 0.1231 × 10^−7^ (according to Shaikh et al., 2020) [19]. Also, the estimated recovery rate (γ) extracted from the data is 0.012 per day. Moreover, the COVID-19 affected family members can survive in the environment from a few hours to several days [3], we considered this value 5 hours in this study and therefore, the virus removal rate (σ) is 0.2/day. We provided the value of the parameters and its sources in Table 1.

Using the above-mentioned values of the parameters, the basic reproduction number was estimated as *R*_0_ = 4.63, with *R*_1_ = 2.42, *R*_2_ = 1.73, *R*_3_ = 0.323 and *R*_4_ = 0.161, establishing the highest contribution of exposed-to-susceptible transmission route to the disease risk.

Next, regarding the impact of undetected cases on the disease risk, table 2 shows a sharp increase in the disease risk with a rise in this number.

## 5. Discussion

This study found a significant contribution of undetected COVID-19 cases to the transmission risk of the disease (*R*_3_ = 0.323). The sensitivity analysis (provided in table 2) showed a rapid increase in the disease risk with the rise in this factor. Also, the highest rate of infection spreading is caused by the exposed individuals ((*R*_1_ = 2.42)), establishing the fact that as the exposed individuals remain asymptomatic in the incubation period of 2–14 days, they can easily transmit the disease unknowingly to the other people in their surroundings. Furthermore, the contaminated environment also contributes to the overall disease risk.

Next, the conceivable implications of this study are specified here. At the current time, when the pandemic claims billions of lives globally with no signs of slowing down due to non-availability of vaccine or definitive treatment, the findings of this study may help in detecting better preventive health policy initiatives in terms of decreasing the speed of social transmission. In this regard, the presently proposed actions like social distancing and the implementation of lockdown appear to be justified to curb the pace of disease transmission. Besides, this model hints, the environmental components like fomites might have some role in disease transmission, therefore, the use of personal protective equipment, like masks, gloves, hand sanitizer seems to be essential.

Regarding the strengths of this paper, this is perhaps the first study on COVID-19 that explored the role of positive non-detected cases on disease transmission in the community, using a mathematical model. Furthermore, the use of recent outbreak data makes the results of this study relevant to the current pandemic scenario. Also, the inclusion of the environment as a component of the mathematical model helps to understand the agent, host, and environment relationship of COVID-19 better.

Despite these strengths, this study has a few weaknesses. Since, the basic reproduction number was estimated by taking some parameter values from other studies and an approximated value of the number of undetected cases, the findings of this model may differ slightly from its actual value.

## 6. Conclusions

The COVID-19 disease risk increases with the growing number of undetected cases. Besides, individuals exposed to SARS-CoV-2 infected individuals can unknowingly spread the infection at a very high rate to those around them. Therefore, to control the disease spread, it is crucial to identify and socially isolate such undetected and exposed cases.

## Data Availability

All data sources are properly cited in the manuscript.

https://www.who.int/docs/default-source/wrindia/situation-report/india-situation-report-8.pdf?sfvrsn=cd671813_2

https://phdmah.maps.arcgis.com/apps/opsdashboard/index.html#/2cc0055832264c5296890745e9ea415c

https://en.wikipedia.org/wiki/COVID-19_pandemic_in_Maharashtra#May

## Authors contribution

The concept, design, analysis, and the manuscript draft were primarily prepared by SS^1^. SS^2^ aided in conception, interpretation and hard editing of the manuscript.

## Ethical clearance

An ethical clearance was not required

